# Determinants of sexual and reproductive empowerment: a cross-national analysis in five Sub-Saharan African countries

**DOI:** 10.1101/2024.07.31.24311299

**Authors:** Caroline Moreau, Celia Karp, Shannon N. Wood, Georges Guiella, Peter Gichangi, Frederick Makumbi, Rosine Mosso, Sani Oumarou, Linnea Zimmerman

## Abstract

Research has shown inconsistent associations between women’s empowerment and sexual and reproductive health (SRH) in sub-Saharan Africa (SSA), pointing to a lack of salient measures in this context. A novel measure provides an opportunity to explore different facets of SRH empowerment in SSA.

**Methods:** We used five national surveys among women 15-49 years conducted between 2019-2020 in Kenya (n=5,504), Uganda (n=2,273), Cote d’Ivoire (n=2,329), Burkina Faso (n=4,135), and Niger (n=2,228). Two measures of contraceptive and sexual empowerment were combined into a four-category SRH empowerment measure to identify the opportunity structures associated with SRH empowerment using multinomial logistic regression modeling.

**Results:** SRH empowerment was lowest in Niger and highest in Kenya. Between 15.0% and 21.7% of participants had more sexual but less contraceptive empowerment than their peers, while the opposite was true for 15.4% to 20.4% of participants. Education increased overall SRH empowerment or sexual empowerment alone in all sites and contraceptive empowerment alone in Kenya. Wealth increased overall SRH empowerment or sexual empowerment alone in three sites. Parity increased overall empowerment or contraceptive empowerment alone in all sites but decreased sexual empowerment alone in Uganda. Finally, healthcare provider contact increased overall and contraceptive empowerment in four sites while family planning media messages increased overall empowerment in two sites.

**Conclusion:** This study confirms the multidimensional nature of SRH empowerment, which varies by country, domain, and by women’s parity and social capital. Longitudinal research is needed to comprehend how women gain or lose SRH empowerment and how empowerment predicts SRH outcomes.

## Introduction

Women’s and girls’ empowerment, generally understood as a process by which individuals exert control over their own destinies by actively challenging gender and power inequities, has gained prominence in policy and programmatic arenas to promote human rights and improve health(1). A growing body of research has shown linkages between women’s empowerment and sexual and reproductive health (SRH)(2–5). Inconsistent evidence, however, particularly in sub-Saharan Africa, has led to calls for greater conceptual development(6) and consideration for contextual variations in the expression of empowerment(7,8). Empowerment results from the interface between *agency* - the ‘ability to make effective choices and to transform these choices into desired outcomes’ - and *opportunity structures* - the sociocultural and economic resources and the political and legal environment that shape women’s choices(9,10). The interplay between these two dimensions is rarely reflected in current research, which either focuses on women’s opportunity structures (e.g., access to education, wealth, or health information and proximity to health services) or agency, such as their freedom of movement or household decision-making(8,11,12). While relevant for understanding specific aspects of empowerment, these proxy measures lack a clear conceptual linkage to SRH behaviors and outcomes(13).

Promising new measures of SRH agency, including those reflecting negotiation skills, autonomy, decision-making, and control over sexual intercourse and contraceptive use, facilitate a broader investigation of the full spectrum of women’s SRH agency and its contribution to health (11,12,14–17). In particular, the recent development and validation of a women and girls’ SRH empowerment index, grounded in the voices of women living in diverse communities across the African continent (Ethiopia, Nigeria, Uganda) (11,12) provides an opportunity to better understand African women’s ability to set and achieve their SRH goals (17). The SRH empowerment measure, which in fact, solely focuses on SRH agency, is not only salient to the sub-Saharan context, but addresses several conceptual limitations of prior empowerment research, by considering SRH agency as a dynamic and multidimensional process that interacts with opportunity structures to inform SRH outcomes. The structure and content of the SRH empowerment measure offers an opportunity to understand how levels of agency vary according to the domain of decisions (e.g., sexual versus contraceptive decisions) and to the nature of decisions (e.g., setting goals versus acting on these goals), Specifically, the measure aligns with the World Bank’s empowerment framework (10), which considers agency as a process moving from existence of choice (i.e., setting goals aligned with one’s values) to exercise of choice (i.e., translation of goals into action), ultimately informing the achievement of choice (i.e., realization of one’s goals). The psychometric properties of the SRH empowerment multidimensional measure were initially validated in a pilot study conducted in three Sub-Saharan countries(17). The measure offers an opportunity to fully explore the empowerment framework, by identifying the opportunity structures that support or constrain each component of SRH empowerment (contraceptive and sexual) as well as the conditions favoring overall SRH empowerment, to inform programmatic efforts.

Using the newly developed SRH empowerment measure, we seek to (1) empirically test the distinction between sexual and contraceptive empowerment, (2) understand the opportunity structures that enable each component, and (3) identify the conditions informing the intersection of sexual and contraceptive empowerment across a diversity of sociocultural settings in sub-Saharan Africa.

## Materials and Methods

### Setting and study samples

This study draws from five population-based surveys conducted in 2019-2020, as part of the Performance Monitoring for Action (PMA) study in Kenya, Uganda, Cote d’Ivoire, Burkina Faso, and Niger, which are the five PMA countries including national samples. These countries cover different geographies in Sub-Saharan Africa (East and West Africa), and represent a diversity of socioeconomic, and cultural backgrounds. According to the United Nation’s human development composite measures, Kenya and Cote d’Ivoire show medium levels of human development, ranking 152^nd^ and 159^th^ respectively on the human development index, while Uganda, Burkina Faso, and Niger show low levels of human development, ranking 166^th^, 184^th^ and 189^th^ respectively(20). These countries also vary according to their gender inequality index, lower in East Africa (Kenya and Uganda rank 128^th^ and 131^st^, respectively), than in West Africa (Niger, Cote d’Ivoire and Burkina Faso rank 153^th^, 155^th^ and 157^th^ respectively(21).

PMA surveys follow a common protocol, including sampling, survey instruments and data collection procedures are described by Zimmerman et al. (22) and further detailed at pmadata.org. Briefly, in each site, a multistage cluster sampling design was used to select a representative sample of the female population aged 15-49 years, who were screened for eligibility and who consented to participate. All participants provided oral consent, and unmarried minors provided parental consent and ascent to participate. Women responded to a face-to-face survey, administered by trained local interviewers, which solicited information on their sociodemographic background, reproductive history, fertility preferences, contraceptive knowledge, and practices, and included a series of questions related to SRH empowerment.

Altogether 3,633 to 9,477 women across geographies completed the survey with participation rates ranging from 95.4% in Niger to 98.7% in Kenya (Table 1). For this analysis, we selected women who were partnered at the time of the survey and ever had sexual intercourse and further restricted to participants who had no missing information about the SRH empowerment measure. Participants were considered to have missing information about SRH empowerment if they were missing half of the items or more on any of the four subscales comprising SRH empowerment, described further below. (Table 1). Our final analytic samples ranged from 2,273 women in Uganda to 5,504 women in Kenya.

**Table 1:** Study samples.

|  | Total female samples | Samples of partnered women who ever had sex. | Analytic samples (partnered women who ever had sex and data on SRH empowerment measures) |
| --- | --- | --- | --- |
| Burkina Faso (2019) | 6,590 | 4,297 | 4,135 |
| Cote d'Ivoire (2020) | 4,135 | 2,499 | 2,329 |
| Niger (2020) | 3,633 | 2,526 | 2,288 |
| Kenya (2019) | 9,477 | 5,541 | 5,504 |
| Uganda (2020) | 3,938 | 2,291 | 2,273 |

This study was approved by ethical review committees at Johns Hopkins School of Public Health (IRB14702; MOD18860; MOD3903; MOD4748), Comité d’Ethique pour la recherche en santé and the Ministère de la Santé et Ministère de l’Enseignement Supérieur, de la Recherche Scientifique et de l’Innovation in Burkina Faso, Comité d’Ethique de la Recherche Institut Pasteur de la Côte d’Ivoire, Kenya Medical Research Institute (KEMRI) Ethics Review Committee in Kenya, Comité Consultatif National d’Ethique in Niger, and Makerere University School of Public Health Higher Degrees, Research and Ethics Committee in Uganda.

### Measures

SRH empowerment is a two-dimensional measure, evaluating the degree to which women make decisions related to sex and contraception and act on these decisions (17). This cross-cultural measure is structured according to the World Bank’s framework, which describes agency as the movement from existence of choice to exercise of choice, ultimately leading to achievement of choice (10). The measure, which centers on existence and exercise of choice as predictors of achievement of choice was developed and tested using a mixed-methods study conducted in Ethiopia, Uganda, and Nigeria (17–19). The 15-item multidimensional construct distinguishing the dimensions of existence and exercise of choice within broader domains of sexual and contraceptive empowerment (17). The measure was restricted to 13 items as one item of contraceptive exercise of choice subscale was accidently skipped and one item from the one item of sexual exercise of choice subscale was dropped after assessing its psychometric properties in eight PMA geographies (Table 2). Internal reliability testing yielded Cronbach alphas ranging from 0.68 to 0.73 for contraceptive empowerment and from 0.67 to 0.77 for sexual empowerment.

**Table 2:** SRH empowerment items by domain and.

| **Sexual empowerment (6 items)** |  |
| --- | --- |
| **Existence of choice** | If I refuse sex with my husband/partner, he may physically hurt me |
|  | If I refuse sex with my husband/partner, he may force me to have sex |
|  | If I refuse sex with my husband/partner, he may stop supporting me |
|  | I am confident I can tell my husband/partner when I want to have sex |
| **Exercise of choice** | I am able to decide when to have sex |
|  | If I do not want to have sex, I can tell my husband |
| **Contraceptive empowerment (7 items)** |  |
| **Existence of choice** | If I use family planning, my husband/partner may seek another sexual partner |
|  | If I use family planning, I may have trouble getting pregnant the next time I want to |
|  | If I use family planning, it may cause conflict in my relationship |
|  | If I use family planning, my body may experience side effects that will disrupt relations |
|  | If I use family planning, my children may not be born normal |
| **Exercise of choice** | I can decide to switch from one family planning method to another if I want to |
|  | I feel confident telling my provider what is important when selecting a method |

We first created two measures reflecting sexual empowerment (six items) and contraceptive empowerment (seven items). Each domain-specific empowerment score was calculated based on women’s responses to each item, with item responses defined on a five-point Likert scale (ranging from strongly agree to strongly disagree). The sexual empowerment scale included three items representing existence of choice and three items representing exercise of choice, while the contraceptive empowerment scale comprised five items reflecting existence of choice and three reflecting exercise of choice (Table 2). For each measure, item responses were averaged separately for each dimension (i.e., existence of choice and exercise of choice), and subsequently averaged across the two dimensions. Missing items ranged from 6%-12% for contraceptive existence of choice, 0.5%- 6.2% for contraceptive exercise of choice and fell below 2.5% for sexual existence and exercise of choice. Missing items were replaced by the average score of their respective dimension for the calculation of each of the four subscales. Final scores ranged from 1-5 for each domain, with higher scores indicating greater empowerment in that domain (i.e., sex, contraception) (17).

Next, we created a relative measure of SRH empowerment reflecting 1) the intersection of the sexual and contraceptive empowerment measures and 2) comparing individuals to their peers. The continuous domain-specific measures of sexual and contraceptive empowerment were dichotomized at the median in each site to create a four-category relative measure of SRH empowerment (lower SRH empowerment, higher sexual empowerment only, higher contraceptive empowerment, and higher SRH empowerment). Respondents who scored below the median on the contraceptive empowerment measure and above the median on the sexual empowerment measure compared to their peers, were considered to have higher than average sexual empowerment only, while those who scored above the median on contraceptive empowerment and below the median on sexual empowerment compared to their peers, were considered to have higher than average contraceptive empowerment only. Respondents who, compared to their peers, consistently fell below or above the median for both measures of sexual and contraceptive empowerment were classified as having lower than average SRH empowerment or higher than average SRH empowerment, respectively. For simplicity we refer to these categories as “lower SRH empowerment”, “higher sexual empowerment only”, “higher contraceptive empowerment only” and “higher SRH empowerment”.

We considered sociodemographic characteristics (age, marital status, education, employment, household wealth), life course experiences (parity), and other resources (recent access to FP messages, cell phone ownership, recent contact with the health system and area of residence (rural/urban)) as the opportunity structures that could enable and enhance sexual and contraceptive empowerment. Recent access to FP messages included any exposure to FP messages via the radio, TV, magazines, or social media in the last 12 months, while recent contact with the health system was defined as having been to a health facility or having been visited by a healthcare worker in the last 12 months.

### Analysis

We first conducted a descriptive analysis, calculating mean and median scores of sexual empowerment and contraceptive empowerment by site. We also examined correlations between domains (sexual empowerment and contraceptive empowerment), between dimensions domains (existence and exercise of choice) across domains and within each domain. Next, we conducted bivariate and multivariate analysis in the form of linear regression models to identify sociodemographic characteristics, life course experiences, and other resources associated with sexual empowerment and contraceptive empowerment. We further examined how women’s combined SRH empowerment (operationalized by the four-categorical relative measure), varied according to women’s opportunity structures (sociodemographic characteristics, life experiences and other resources) using bivariate and multivariate multinomial regression models. Analyses were stratified by site. They were also weighted and accounted for clustering to reflect the complex survey design. All analyses were conducted using Stata 17.

## Results

The characteristics of the women in our sample, by site, are displayed in Table 3. The mean age of respondents varied from 29.0 years in Niger to 32.3 years in Kenya (data not shown). Marital status and parity varied widely across countries, with a higher proportion of non-married, partnered women in Uganda and Cote d’Ivoire, and higher proportion of low-parity women in Kenya and Cote d’Ivoire. Likewise, while most women had no education in the three West African countries, a majority attended at least primary education in Uganda and Kenya and 31% and 42.6%, respectively, had a secondary education. Women in Cote d’Ivoire and Niger were least likely to work outside of the home, while employment reached 64% in Uganda. Between 63% to 82% of women had contact with the healthcare system in the last 12 months (through facilities or community health workers) and between 49.7% of women in Cote d’Ivoire and 86.2% in Kenya had heard or viewed family planning messages on the radio, TV, social media, or magazines in the last 12 months.

**Table 3:** Distribution of women’s opportunity structures by site.

|  |  | Burkina Faso | Cote d'Ivoire | Niger | Kenya | Uganda |
| --- | --- | --- | --- | --- | --- | --- |
| Age | 15-24 | 26.0% | 20.7% | 30.5% | 19.2% | 27.7% |
|  | 25-34 | 37.6% | 41.6% | 41.5% | 43.1% | 40.3% |
|  | 35-49 | 36.5% | 37.7% | 28.0% | 37.7% | 32.0% |
| Marital status | Not married/cohabitating | 9.7% | 38.4% | 0.7% | 9.8% | 56.0% |
|  | Married | 90.3% | 61.6% | 99.3% | 90.2% | 44.0% |
| Parity | 0-2 children | 36.0% | 43.7% | 34.0% | 41.0% | 38.5% |
|  | 3-4 children | 27.0% | 28.7% | 27.6% | 32.8% | 25.9% |
|  | 5+ children | 37.0% | 27.5% | 38.4% | 26.1% | 35.6% |
| Education | None | 70.7% | 52.4% | 75.2% | 6.1% | 8.2% |
|  | Primary | 17.1% | 27.1% | 14.3% | 51.3% | 60.7% |
|  | Secondary | 12.1% | 20.5% | 10.5% | 42.6% | 31.1% |
| Employment in the last 12 months | No | 45.2% | 68.5% | 78.3% | 37.5% | 35.9% |
|  | Yes | 54.8% | 31.5% | 21.8% | 62.5% | 64.1% |
|  | No | 6.9% | 7.3% | 35.5% | 12.5% | 19.6% |
| Has a mobile phone | Yes | 90.1% | 92.7% | 64.5% | 87.5% | 80.4% |
| Residence | Rural | 82.6% | 47.1% | 85.2% | 71.1% | 71.2% |
|  | Urban | 17.4% | 52.9% | 14.8% | 28.9% | 28.8% |
| Contact with health services | No | 18.0% | 26.2% | 36.8% | 26.9% | 26.2% |
|  | Yes | 82.0% | 73.8% | 63.2% | 73.1% | 73.8% |
| Media exposure to FP message in the last 12 months | No | 34.9% | 50.3% | 53.6% | 13.8% | 49.7% |
|  | Yes | 65.1% | 49.7% | 46.4% | 86.2% | 50.3% |

### Sexual and contraceptive empowerment scores

Figure 1 presents mean sexual, contraceptive, and overall SRH empowerment scores by site. Mean empowerment scores for both domains were lowest in Niger and highest in Kenya, with sexual empowerment ranging from 3.35 to 3.98 and contraceptive empowerment scores varying from 3.68 to 4.24. In all sites, mean contraceptive empowerment scores were higher than mean sexual empowerment scores, with the greatest difference observed in Burkina Faso (0.55 points) and the lowest difference in Côte d’Ivoire (0.18 points).

**Figure 1:**
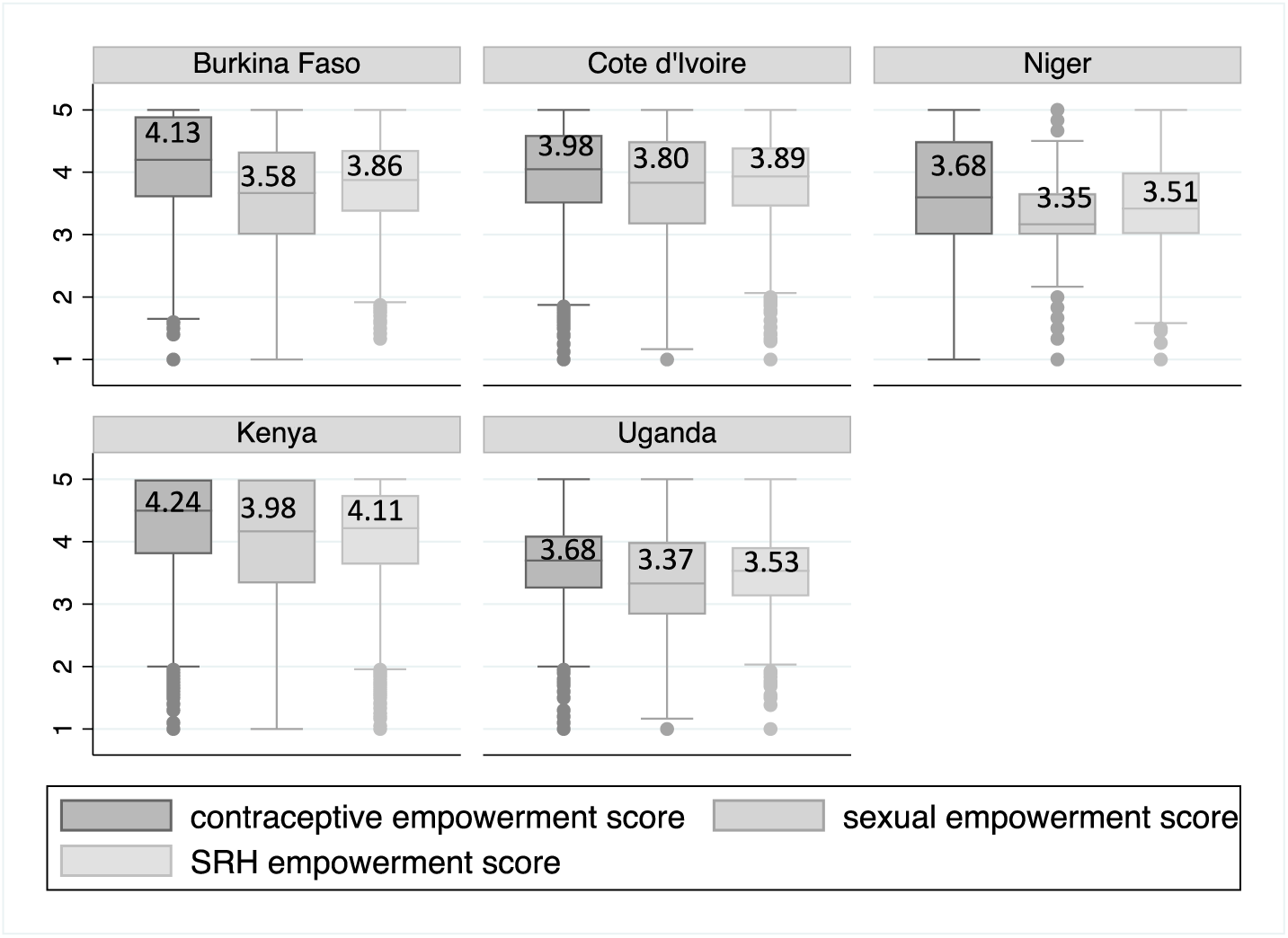
Mean sexual and contraceptive empowerment and overall SRH-empowerment scores by site.

While differences in sexual and contraceptive empowerment scores were small, the measures were moderately correlated in all countries but Kenya, with correlation coefficients ranging from 0.25 to 0.29 (Figure 2). Contraceptive and sexual empowerment scores were more strongly correlated in Kenya (correlation=0.42) (Figure 2). A more detailed comparison of domain and dimension-specific measures indicates low correlations between sexual and contraceptive existence of choice (ranging from a correlation coefficient of 0.28 in Uganda to 0.42 in Niger) and between sexual and contraceptive exercise of choice (from 0.18 in Uganda to 0.30 in Niger) (Appendix 1). Additionally, within each domain, existence and exercise of choice were poorly correlated ranging from -0.17 in Niger to 0.22 in Kenya for existence and exercise of sexual choice and from 0.07 in Uganda to 0.18 in Kenya for existence and exercise of contraceptive choice (Appendix 1).

**Figure 2:**
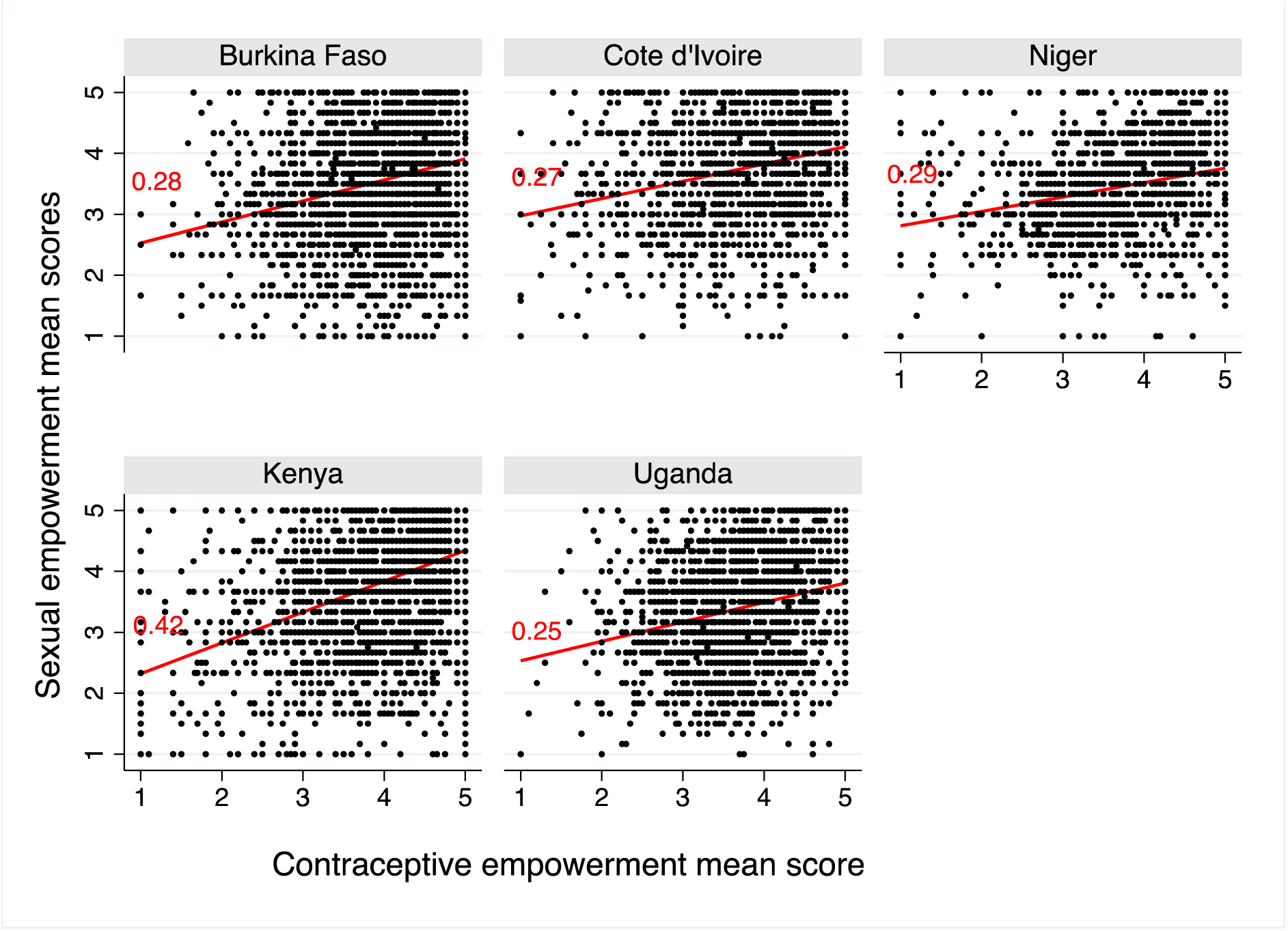
Correlation between sexual and contraceptive empowerment mean scores by site.

Bivariate analysis of the opportunity structures that could enhance sexual and contraceptive empowerment, respectively, show these factors vary by country and by empowerment domain (Table 4). Across sites, education and wealth were generally associated with increased sexual empowerment and, to a lesser extent, with contraceptive empowerment. Parity was associated with increased contraceptive empowerment in two Western African sites (Burkina Faso and Cote d’Ivoire) but correlated with decreased sexual empowerment in all sites except Niger. In all sites, women who had been in recent contact with a provider/community health worker or had heard FP messages in the media exerted higher contraceptive empowerment, and to a lesser degree, higher sexual empowerment. Likewise, cell phone owners reported greater sexual and contraceptive empowerment in three of the five sites. Other factors varied more across sites. For instance, urban residence was associated with higher sexual empowerment in three sites but with contraceptive empowerment in only one site. Likewise, sexual empowerment increased with age in Cote d’Ivoire, but decreased with age in Uganda, while age was unrelated to contraceptive empowerment in all sites.

**Table 4:** Factors associated with sexual empowerment and contraceptive empowerment (FP): bivariate analysis.

|  |  | Burkina Faso |  | Cote d'Ivoire |  | Niger |  | Kenya |  | Uganda |  |
| --- | --- | --- | --- | --- | --- | --- | --- | --- | --- | --- | --- |
|  |  | Sex power | FP power | Sex power | FP power | Sex power | FP power | Sex power | FP power | Sex power | FP power |
| Age | 15-24 | 3.56 | 4.06 | **3.66** | 3.92 | 3.35 | 3.68 | 4.03 | 4.27 | **3.51** | 3.65 |
|  | 25-34 | 3.62 | 4.16 | **3.83** | 4.03 | 3.36 | 3.72 | 3.99 | 4.26 | **3.41** | 3.69 |
|  | 35-49 | 3.56 | 4.15 | **3.83** | 3.96 | 3.33 | 3.61 | 3.95 | 4.22 | **3.22** | 3.70 |
| Marital status | Not married | 3.44 | 4.07 | **3.97** | 4.00 |  |  | 4.11 | 4.24 | 3.39 | 3.69 |
|  | Married/cohabitating | 3.60 | 4.14 | **3.69** | 3.97 |  |  | 3.97 | 4.28 | 3.35 | 3.67 |
| Parity | 0-2 children | **3.61** | **4.05** | **3.86** | **3.93** | 3.35 | 3.62 | **4.11** | **4.31** | **3.53** | 3.63 |
|  | 3-4 children | **3.63** | **4.17** | **3.85** | **4.07** | 3.38 | 3.74 | **3.97** | **4.29** | **3.37** | 3.76 |
|  | 5+ children | **3.52** | **4.18** | **3.62** | **3.95** | 3.32 | 3.68 | **3.8** | **4.09** | **3.21** | 3.67 |
| Education | No education | **3.54** | 4.13 | **3.57** | **3.91** | **3.29** | **3.64** | **3.63** | **3.75** | **3.11** | 3.51 |
|  | Primary | **3.52** | 4.11 | **3.92** | **3.98** | **3.45** | **3.73** | **4.86** | **4.20** | **3.27** | 3.65 |
|  | Secondary | **3.89** | 4.19 | **4.20** | **4.16** | **3.60** | **3.84** | **4.18** | **4.37** | **3.65** | 3.79 |
| Wealth tertile | Lowest | 3.61 | 4.13 | **3.48** | **3.78** | **3.21** | 3.55 | **3.76** | **4.08** | **3.22** | 3.60 |
|  |  |  |  | **3.75** | **3.95** |  |  | **3.93** | **4.23** | **3.30** | 3.65 |
|  | Middle | 3.53 | 4.16 | **3.89** | **4.08** | **3.32** | 3.68 | **3.97** | **4.27** | **3.25** | 3.70 |
|  |  |  |  | **3.84** | **4.04** |  |  | **4.07** | **4.33** | **3.46** | 3.67 |
|  | Highest | 3.62 | 4.10 | **4.05** | **4.07** | **3.51** | 3.8 | **4.27** | **4.35** | **3.68** | 3.79 |
| Phone ownership | Yes | 3.42 | 4.01 | 3.81 | 3.98 | **3.39** | **3.76** | **4.02** | **4.27** | **3.40** | **3.70** |
|  | No | 3.60 | 4.14 | 3.66 | 3.92 | **3.28** | **3.52** | **3.70** | **4.03** | **3.27** | **3.58** |
| Employment | Yes | 3.49 | 4.14 | 3.76 | 3.97 | 3.40 | 3.74 | 4.01 | **4.29** | 3.38 | **3.71** |
|  | No | 3.70 | 4.12 | 3.88 | 4.01 | 3.33 | 3.66 | 3.93 | **4.16** | 3.37 | **3.63** |
| Residence | Rural | **3.65** | 4.06 | 3.72 | 3.93 | **3.30** | **3.63** | **3.93** | 4.22 | 3.28 | 3.67 |
|  | Urban | **3.82** | 4.01 | 3.86 | 4.02 | **3.60** | **4.00** | **4.11** | 4.30 | 3.63 | 3.72 |
| Contact with health provider | Yes | 3.57 | **4.16** | **3.83** | **4.03** | **3.44** | **3.79** | 4.00 | **4.30** | **3.40** | **3.71** |
|  | No | 3.62 | **4.01** | **3.71** | **3.84** | **3.19** | **3.48** | 3.92 | **4.10** | **3.29** | **3.56** |
| Media exposure to FP message in the last 12 | yes | 3.64 | **4.18** | **3.94** | **4.05** | **3.4** | **3.76** | 4.03 | **4.28** | **3.41** | **3.7** |
|  | no | 3.48 | **4.05** | **3.66** | **3.91** | 3.30 | **3.61** | **3.70** | **3.99** | **3.20** | **3.58** |

**bold numbers are indicative of significant associations $p < 0.05$ .**

In multivariate analysis (Table 5), associations with education were mostly maintained, while those between wealth and sexual empowerment and contraceptive empowerment remained significant in three sites (Cote d’Ivoire, Niger, and Kenya) and two sites (Burkina Faso and Cote d’Ivoire), respectively. Parity was still positively associated with contraceptive empowerment in two sites (Burkina Faso and Uganda) and decreased sexual empowerment in one site (Kenya). Finally recent contact with the health care system was associated with increased contraceptive empowerment in all sites but Uganda, and sexual empowerment in one site (Niger), while FP media exposure was associated with increased sexual empowerment and contraceptive empowerment in two different sites.

**Table 5:**
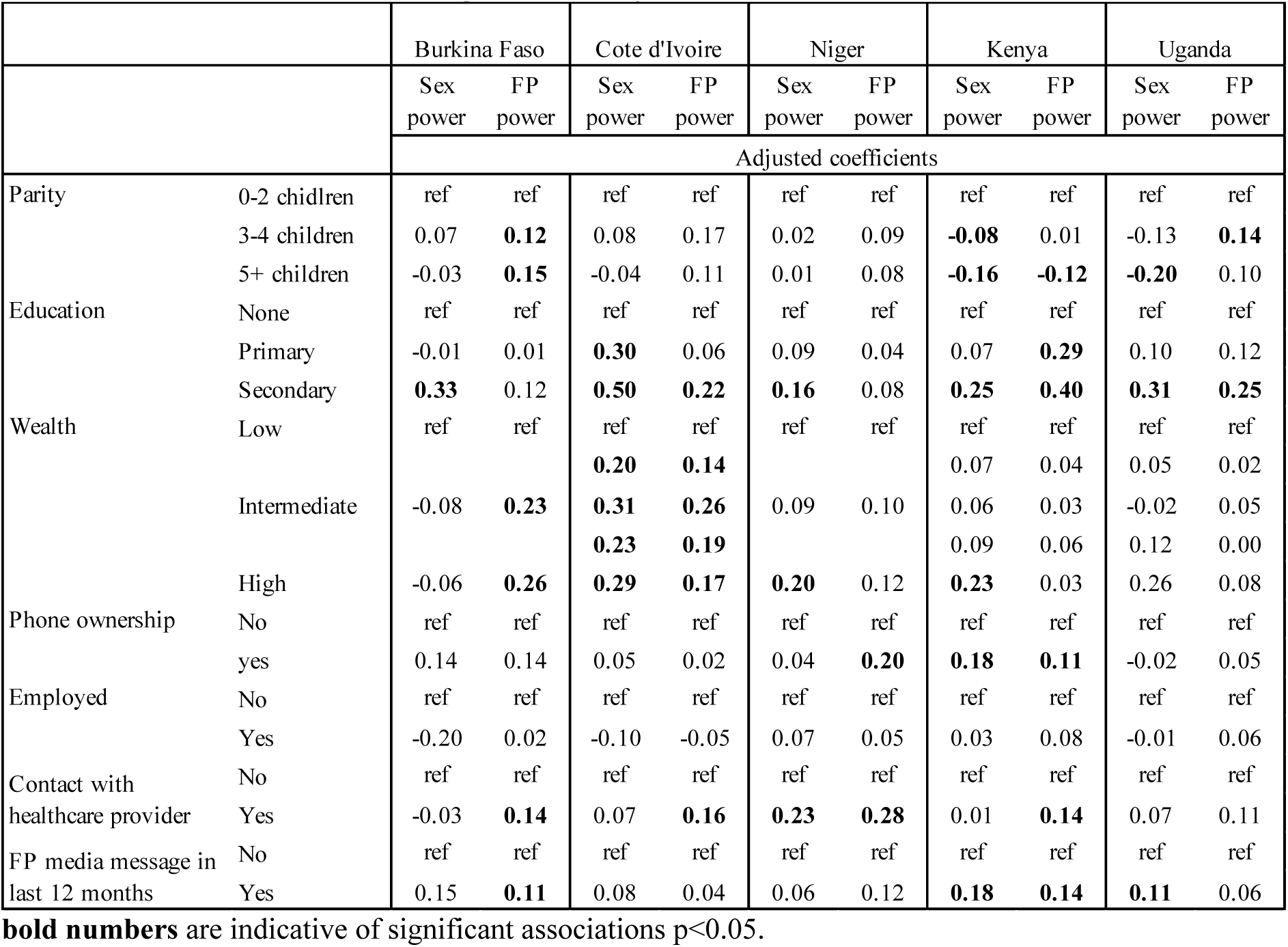
Factors associated with sexual empowerment and contraceptive empowerment (FP): results from multivariate linear regression analysis.

|  |  | Burkina Faso |  | Cote d'Ivoire |  | Niger |  | Kenya |  | Uganda |  |
| --- | --- | --- | --- | --- | --- | --- | --- | --- | --- | --- | --- |
|  |  | Sex power | FP power | Sex power | FP power | Sex power | FP power | Sex power | FP power | Sex power | FP power |
|  |  | Adjusted coefficients |  |  |  |  |  |  |  |  |  |
| Parity | 0-2 children | ref | ref | ref | ref | ref | ref | ref | ref | ref | ref |
|  | 3-4 children | 0.07 | **0.12** | 0.08 | 0.17 | 0.02 | 0.09 | **-0.08** | 0.01 | -0.13 | **0.14** |
|  | 5+ children | -0.03 | **0.15** | -0.04 | 0.11 | 0.01 | 0.08 | **-0.16** | **-0.12** | **-0.20** | 0.10 |
| Education | None | ref | ref | ref | ref | ref | ref | ref | ref | ref | ref |
|  | Primary | -0.01 | 0.01 | **0.30** | 0.06 | 0.09 | 0.04 | 0.07 | **0.29** | 0.10 | 0.12 |
|  | Secondary | **0.33** | 0.12 | **0.50** | **0.22** | **0.16** | 0.08 | **0.25** | **0.40** | **0.31** | **0.25** |
| Wealth | Low | ref | ref | ref | ref | ref | ref | ref | ref | ref | ref |
|  |  |  |  | **0.20** | **0.14** |  |  | 0.07 | 0.04 | 0.05 | 0.02 |
|  | Intermediate | -0.08 | **0.23** | **0.31** | **0.26** | 0.09 | 0.10 | 0.06 | 0.03 | -0.02 | 0.05 |
|  |  |  |  | **0.23** | **0.19** |  |  | 0.09 | 0.06 | 0.12 | 0.00 |
|  | High | -0.06 | **0.26** | **0.29** | **0.17** | **0.20** | 0.12 | **0.23** | 0.03 | 0.26 | 0.08 |
| Phone ownership | No | ref | ref | ref | ref | ref | ref | ref | ref | ref | ref |
|  | yes | 0.14 | 0.14 | 0.05 | 0.02 | 0.04 | **0.20** | **0.18** | **0.11** | -0.02 | 0.05 |
| Employed | No | ref | ref | ref | ref | ref | ref | ref | ref | ref | ref |
|  | Yes | -0.20 | 0.02 | -0.10 | -0.05 | 0.07 | 0.05 | 0.03 | 0.08 | -0.01 | 0.06 |
| Contact with healthcare provider | No | ref | ref | ref | ref | ref | ref | ref | ref | ref | ref |
|  | Yes | -0.03 | **0.14** | 0.07 | **0.16** | **0.23** | **0.28** | 0.01 | **0.14** | 0.07 | 0.11 |
| FP media message in last 12 months | No | ref | ref | ref | ref | ref | ref | ref | ref | ref | ref |
|  | Yes | 0.15 | **0.11** | 0.08 | 0.04 | 0.06 | 0.12 | **0.18** | **0.14** | **0.11** | 0.06 |

**bold numbers** are indicative of significant associations $p < 0.05$ .

The intersection of sexual and contraceptive empowerment, depicted in Figure 3, shows that high SRH empowerment, where women are in the highest median for both sexual and contraceptive empowerment, ranges from 23.3% of women in Niger to 33.9% in Kenya. In contrast, 30.7% of women in Uganda to 42.3% of women in Niger had consistently lower SRH empowerment, scoring below the median for both empowerment subscales, compared to their peers. Between 31.9% of women in Kenya and 41.9% of women in Uganda scored above the median in one domain of empowerment and below the median in the other domain, compared to their peers. Specifically, 15.0% of women in Kenya to 21.7% in Uganda had more sexual empowerment but less contraceptive empowerment than their peers, while the opposite was true among 15.4% of women in Niger and up to 20.4% of women in Côte d’Ivoire. Differences in mean scores between sexual and contraceptive empowerment measures averaged 0.83 and 1.61 points (on a scale from 1-5) among respondents who had only sexual empowerment or only contraceptive empowerment, while the difference was reduced to 0.40 and 0.17 points among respondents who had consistently low or high empowerment across the two dimensions.

**Figure 3:**
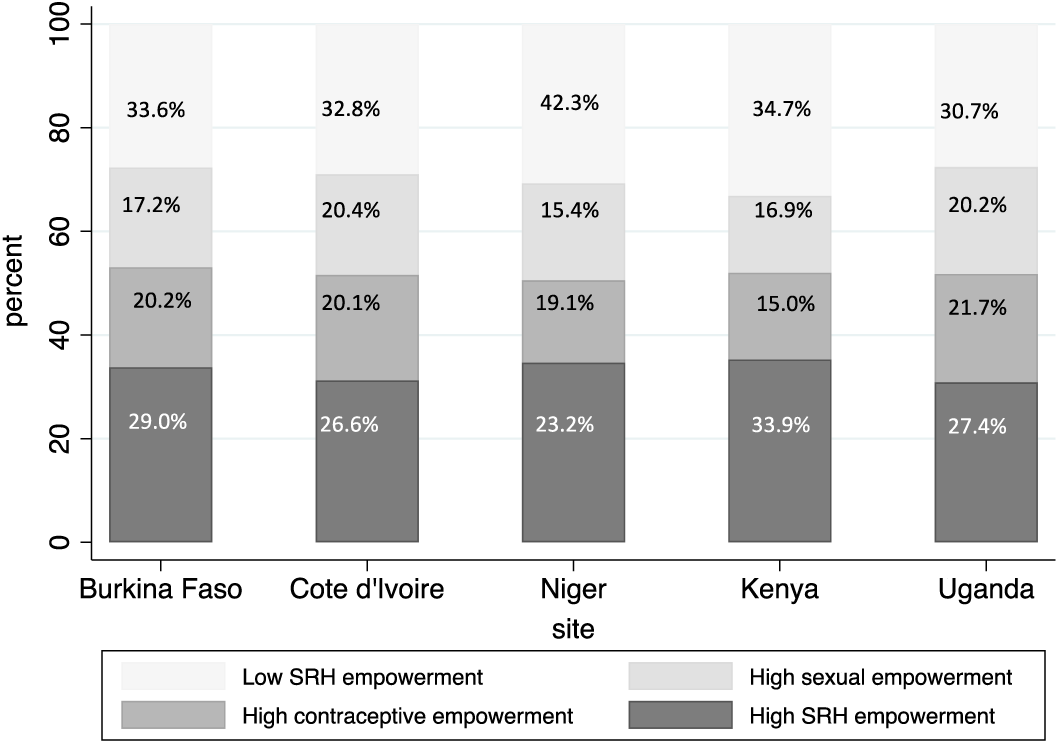
SRH empowerment by site

Patterns of relative SRH empowerment, as a combination of sexual and contraceptive empowerment, varied according to women’s reproductive life cycle, in addition to their social resources (Table 6). In three sites (Burkina Faso, Cote d’Ivoire, and Uganda), increasing parity was associated with a greater percentage of women with higher contraceptive empowerment alone but a lower percentage of women with higher sexual empowerment alone. In West African countries, these opposing effects resulted in little variation in the percentages of women with overall higher or lower SRH empowerment by parity, while in the two Eastern African sites, increasing parity correlated with a rise in the percentages of women with lower overall SRH empowerment and a decline in the percentages of women with higher overall SRH empowerment. In all sites, as education increased, the percentage of women who had higher SRH empowerment increased, while the percentage who had lower SRH empowerment decreased (although non-significant in Uganda, p=0.11). In addition, the percentage of women with higher sexual empowerment alone increased while the percentage of women with higher contraceptive empowerment alone dropped with education in all sites but Kenya (and Niger in the case of contraceptive empowerment). While other factors varied by site, notable associations included increasing percentages of women with higher SRH empowerment and decreasing percentages of women with high lower SRH empowerment with wealth and cell phone ownership in Niger and Kenya. Similar patterns of overall SRH empowerment variations were also observed among women with recent contact with the healthcare system in Cote d’Ivoire, Niger, and Kenya as well as among women exposed to FP messages in Cote d’Ivoire and Kenya.

**Table 6:**
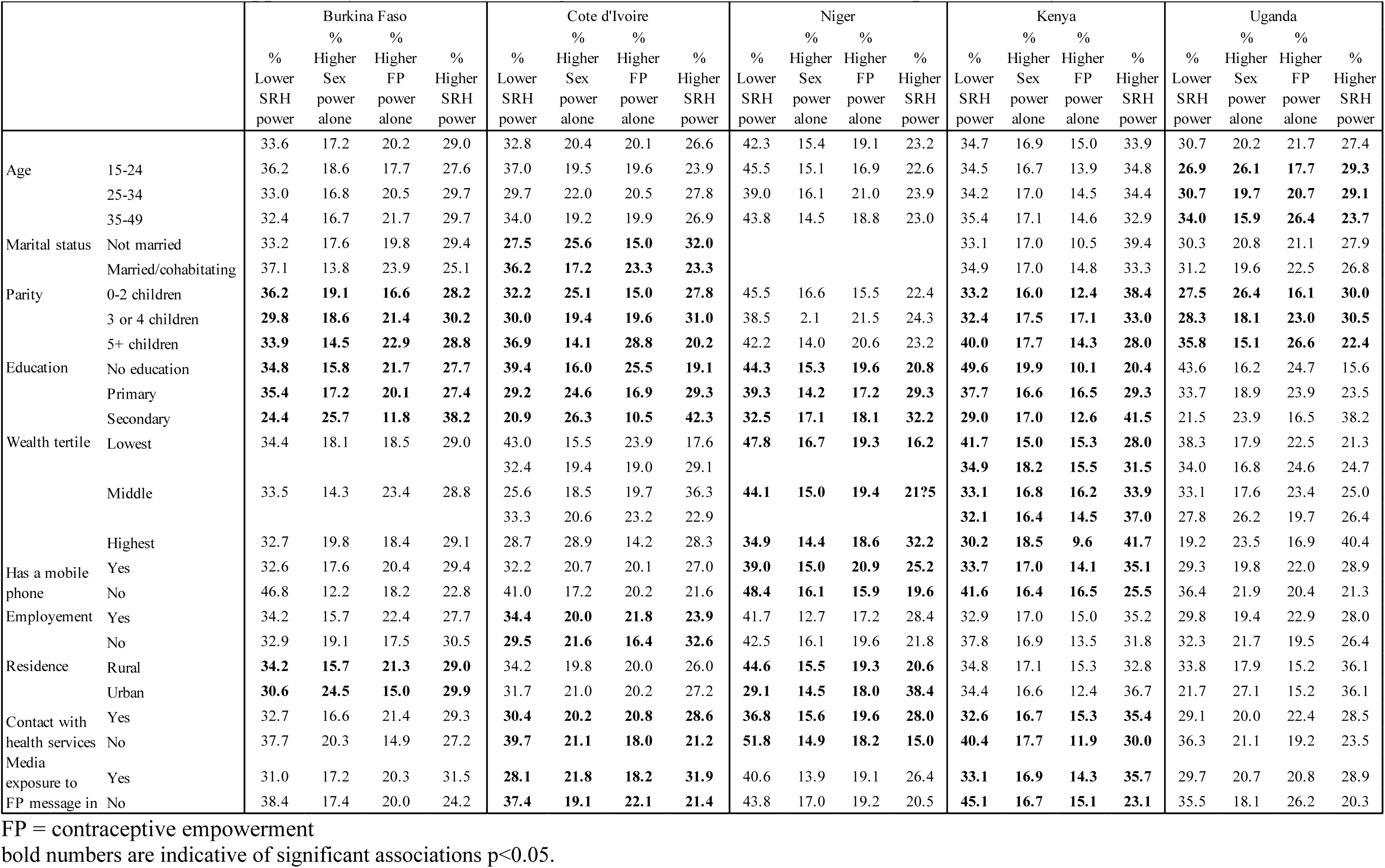
Distribution of opportunity structures according to the relative measure of SRH empowerment, by site.

In multivariate analysis, the role of parity and education remained consistent in most sites, while wealth remained associated with increases in the relative risk ratio of higher SRH empowerment or sexual empowerment alone in three sites (Cote d’Ivoire, Kenya, Uganda). Recent contact with the healthcare system was related to increased relative risk ratio of higher overall SRH empowerment or higher contraceptive empowerment in four sites, while exposure to FP messages through the media was associated with higher overall SRH empowerment in two sites (Cote d’Ivoire and Kenya) (Table 7).

**Table 7:**
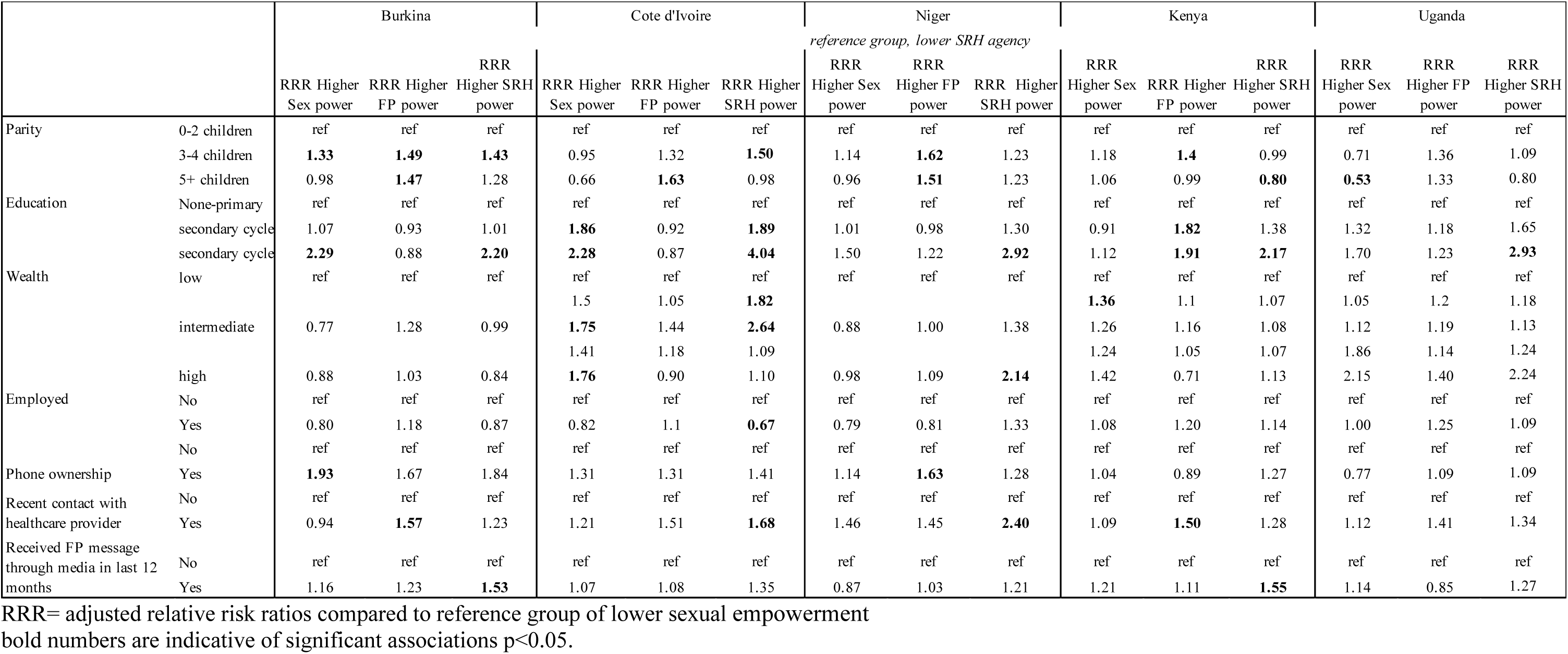
Opportunity structures associated with the relative measure SRH empowerment by site: results from multivariate multinomial regression models.

## Discussion

This cross-national exploration of women’s SRH empowerment in five sub-Saharan countries provides empirical evidence supporting the multidimensional nature of women’s SRH empowerment, which lies at the intersection of agency and opportunity structures and varies according to the nature of decisions considered (sex versus contraception). Using a newly validated measure of SRH empowerment, we found empowerment varied by country and domain, and according to women’s reproductive life cycle and social resources.

Across domains of sex and contraception, empowerment was consistently lowest in Niger and Uganda and highest in Kenya. These differences align with some global indicators of development, such as the United Nation’s human development index(20), offering empirical support for the UN’s agenda of promoting women’s empowerment to accelerate sustainable human development, although directionality of associations still need to be established. However, patterns of sexual and contraceptive empowerment are less aligned with country level indicators of gender equality and SRH, as the gender inequality index is more favorable in Uganda than any our Western African countries(23) while SRH empowerment was more favorable in Cote d’Ivoire and Burkina Faso than Uganda. While the gender inequality index includes dimensions of reproductive health and empowerment along with labor market, these dimensions focus on different aspects of empowerment (education and parliament representation) or reproductive health (maternal mortality and adolescent birth rates)(23), which likely explains differences in the patterns observed. As such, our measure of SRH empowerment may provide complementary information towards monitoring progress towards achieving SDG 5 and SG3 goals.

The study also confirms theoretical perspectives on empowerment, which lies at the intersection of agency and opportunity structures and is multidimensional, varying according to the nature of life decisions. In our study, the low correlation between sexual and contraceptive empowerment scores suggests that women who have power over sexual decisions do not necessarily control contraceptive decisions, and vice versa. These results have both research and programmatic implications, highlighting the need to align empowerment indicators with the SRH outcomes of interest. For example, efforts to promote contraceptive empowerment and informed choice to use contraception may bear little impact on women’s sexual empowerment within their partnerships. Similarly, programs aimed at increasing sexual empowerment and/or reducing sexual violence, may not advance outcomes related to contraceptive use. To simultaneously empower women and girls in achieving their SRH outcomes more holistically, programs must include components promoting both sexual and contraceptive empowerment. As such, contextually validated measures, such as our SRH empowerment index, or other indicators of contraceptive or sexual autonomy, should be prioritized over more traditional measures, such as household decision-making or mobility, that are loosely and inconsistently associated with SRH outcomes in sub-Saharan Africa (6).

The higher levels of contraceptive empowerment compared to sexual empowerment in all countries, could reflect the division of gender roles and power, promoting men’s sexual prowess, while stigmatizing women’s sexuality and instead assigning women the responsibility of regulating their fertility (24). These divisions amplify over the reproductive life course, as we found women who had more children gained contraceptive empowerment in some sites but lost sexual empowerment in the two East African settings. While the directionality of associations cannot be established with cross-sectional data, we hypothesize the observed patterns could be reinforced by the health system, which seems to be associated with increased contraceptive empowerment, but not sexual empowerment, calling for greater programmatic efforts to address women’s SRH needs more holistically. However, these efforts are unlikely to succeed unless they also engage men to challenge gender-specific norms and attitudes surrounding sex, and improve their competencies in preventing sexual risks related to STIs and unintended pregnancy(25).

Our study also explores the theoretical distinction between agency and opportunity structures, showing how different forms of social capital enable different forms of empowerment. We found women’s education and wealth were consistently related to sexual empowerment and to a lesser extent contraceptive empowerment, while employment was not related to either. Education was specifically correlated with contraceptive empowerment in settings where women accessed higher education but not in countries where 70% of the population or more had little to no education. These results confirm the role of education in promoting women’s sexual and contraceptive empowerment (26), while calling for greater understanding of the relationship between women’s economic empowerment and SRH empowerment. Prior studies have raised the alarm on the potential backlash of women’s economic autonomy on their social safety, including exposure to intimate partner violence(27). The intersection of economic and SRH empowerment was beyond the scope of the present study, but merits further exploration given the lack of cross-sectional association between women’s employment and their level of SRH empowerment.

While the current study uses cross-sectional data that prevent the exploration of SRH empowerment as a dynamic process that unfolds from existence to exercise of choices leading to desired outcomes, our results provide support for future longitudinal investigation of this process, given the distinct constructs of existence and exercise of sexual or contraceptive choices described in this study. Specifically, longitudinal studies should examine how SRH empowerment develops from existence to exercise of choice and how each component contributes to future desired SRH outcomes. Our current multidimensional SRH empowerment measure allows such an exploration using repeated measures over time.

Beyond the cross-sectional nature of the study that prevents any causal inferences, our study should be interpreted with several limitations in mind. The SRH empowerment index was missing one item related to exercise of contraceptive choices which likely affects the quality measure. The item was reintroduced in Phase 2, but we could not use Phase 2 data as sexual empowerment was only collected in Phases 1 and 3 of the PMA surveys to reduce survey burden. Additionally, while the measure captures different components of SRH empowerment, some domains are missing, including pregnancy-related empowerment or the ability to determine the timing and outcome of a pregnancy. This domain was initially included in the formative measurement work, but no consistent indicator of pregnancy empowerment emerged across settings (17). Further measurement advances are needed to integrate women’s motivations for childbearing and their ability to act on them, which is distinct from contraceptive empowerment, given the significant gap between pregnancy intentions and contraceptive intentions (28). Another study limitation is the lack of partner perspective, providing little context in which women make SRH decisions, despite the dyadic nature of sexual and reproductive behaviors (29) and the power dynamics informing these decisions (30).

Despite, these limitations, we believe our results contribute important empirical evidence of the multidimensional nature of SRH empowerment and its connection to opportunity structures, pointing to the evolving nature of empowerment over the reproductive life course and the importance of social capital (education) in strengthening women’s ability to set and act on their sexual desires and contraceptive decisions. Longitudinal research is needed to further understand how women gain or lose power over these decisions and how the intersection of agency and opportunity structures, which together constitute empowerment, predict future sexual and contraceptive behaviors.

## Data Availability

The data used for this analysis are publicly available at https://pma.ipums.org/pma/ and https://www.pmadata.org/data/request-access-datasets

https://www.pmadata.org/data/request-access-datasets

## Acknowledgements

The authors would like to thank the women who graciously agreed to share their experiences and perspectives by responding the PMA survey. We also thank Performance Monitoring for Action (PMA) teams, including data collectors, field supervisors, and research staff and for their invaluable work in fielding the PMA surveys. Their involvement made a critical contribution to the quality of the research presented.

## Appendix 1: Correlations between different domains and dimensions of SRH empowerment

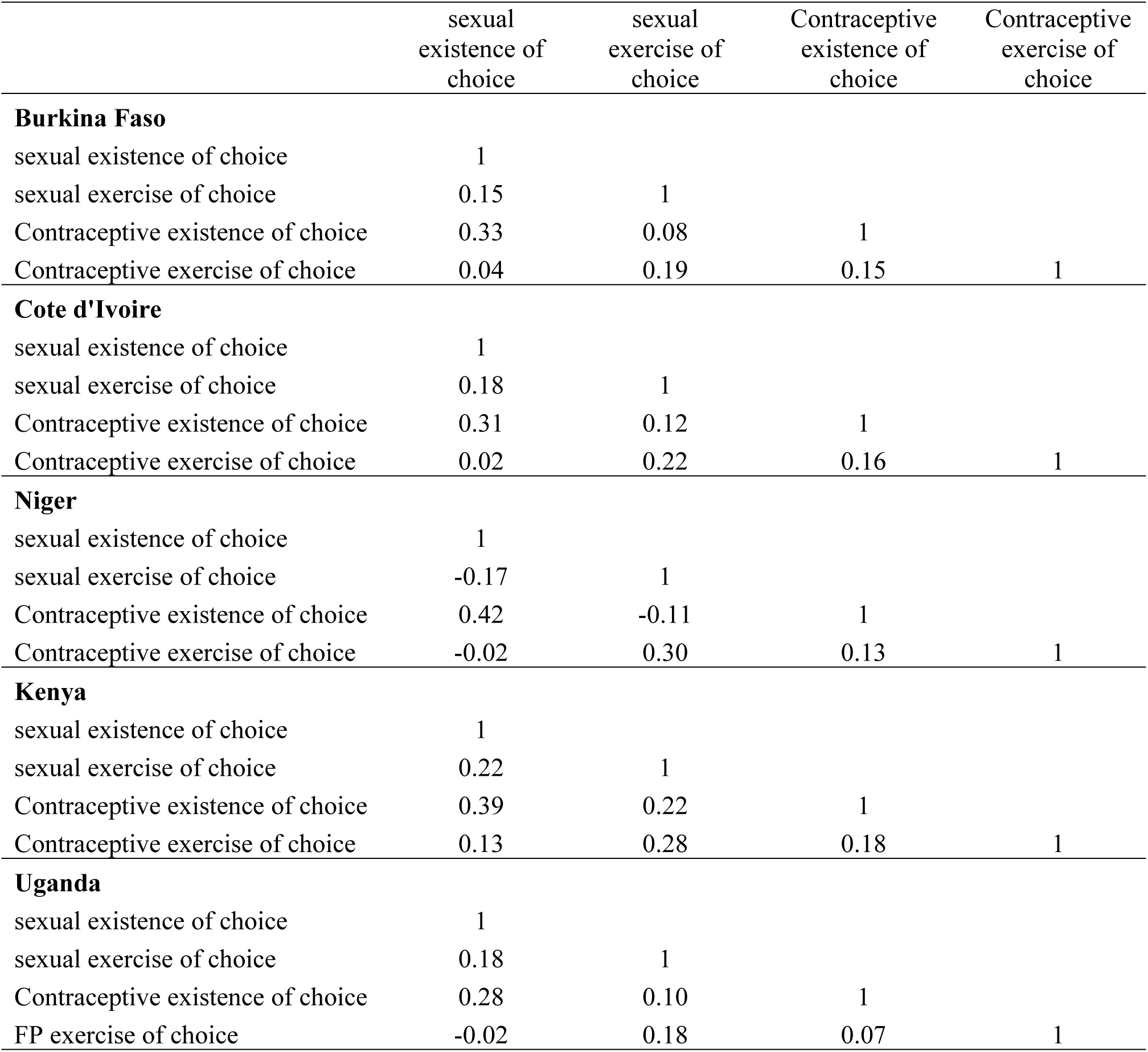

